# Digital Data Collection Reduces Terminal Digit Bias in Haemodynamic and Physical Activity Measures in Pulmonary Arterial Hypertension

**DOI:** 10.1101/2025.05.21.25328105

**Authors:** Alexander M.K. Rothman, James Wason, Mark Toshner

## Abstract

Pulmonary arterial hypertension (PAH) is a progressive condition requiring precise measurement of pulmonary artery pressure (PAP), cardiac output (CO), and exercise capacity for diagnosis and therapeutic evaluation. Terminal digit preference, a form of observer bias favouring values ending in 0 and 5, has been reported in PAH clinical trials, potentially introducing significant measurement error. In the FIT-PH study (NCT04078243), we evaluated whether terminal digit preference occurs in remotely collected cardiopulmonary data from implanted monitors in 35 patients with PAH. Over 18,000 haemodynamic and 34,000 heart rate and activity measurements were analysed. No evidence of terminal digit preference was found in systolic or mean PAP, CO, heart rate, or physical activity. A non-uniform terminal digit distribution in diastolic PAP was observed but attributed to physiological constraints rather than rounding bias. These findings demonstrate that digital remote monitoring minimises observer-related bias, enhancing accuracy in PAH research and clinical care

## Introduction

Pulmonary arterial hypertension (PAH) is a rare progressive disease with significant morbidity.^1^ Randomised clinical trials of PAH therapies use endpoints including cardiac output, pulmonary arterial pressure, and 6-minute walk distance.^2^ A terminal digit preference for values of 0 and 5 for pulmonary artery pressure and 6-minute walk distance has been demonstrated in phase III clinical trials of PAH therapies. ^3^ This bias has the potential to introduce inaccuracies in diagnosis and the assessment of the efficacy of therapeutic interventions. Technological advances provide the capacity to remotely measure cardiopulmonary haemodynamics and physical activity, daily from a patient’s home, using minimally invasive, implanted devices. We evaluated whether terminal digit preference affected remote measurements of heart rate, physical activity, cardiac output, and pulmonary artery pressure in patients with pulmonary arterial hypertension with implanted devices.

## Methods

Following enrolment in Feasibility of Novel Clinical Trial Infrastructure, Design and Technology for Early Phase Studies in Patients with Pulmonary Hypertension (FIT-PH, NCT04078243, REC 19/YH/0354), patients underwent baseline 6-minute walk distance and right heart catheterisation, and implantation of a pulmonary artery pressure monitor and insertable cardiac monitor. Daily, remote measures of heart rate, physical activity, cardiac output, and pulmonary artery pressure were relayed to the clinical care team through secure online portals. Assuming a uniform distribution of terminal digits, each digit (0–9) should be represented at 10%. Deviation from this distribution was tested for systolic, diastolic, and mean pulmonary artery pressure, cardiac output and physical activity using a χ^2^ test. Analyses used SPSS version 29 for macOS (IBM).

## Results

Thirty-five patients underwent right heart catheterisation with implantation of minimally invasive remote monitoring devices (Table 1). Following implantation, 18,646 independent measures of pap and cardiac output and 34,611 independent measures of heart rate, heart rate variability and physical activity were made between November 2019 and May 2023. The distribution of terminal digits for systolic and mean pulmonary artery pressure, cardiac output and physical activity did not deviate from the expected representation (Figure 1A-G). The terminal digits for measures of diastolic pulmonary artery pressure were not equally represented (Figure 1H). The digits 7, 8 and 9 were significantly overrepresented.

**Table 1:** Baseline patient characteristics.

| Demographics | N=35 |
| --- | --- |
| Age (years, mean ± SD) | 51 (28.5) |
| Gender (n, % female) | 25 (71.4) |
| Ethnicity (n, %) |  |
| • White | 33 (94.3) |
| • Asian | 2 (5.7) |
| BMI (mean ±SD) | 27.9 (5.7) |
| ISWT (mean ±SD) | 362.9 (231.7) |
| WHO Functional Class I/II/III/IV (n, %) | 0 /1 (2.9) /29 (82.9) /5 (14.3) |
| NT-proBNP (mean ±SD) | 666.2 (1065.5) |
| PH sub-diagnosis (n, %) |  |
| • IPAH | 28 (80) |
| • HPAH | 1 (2.9) |
| • CTD | 4 (11.4) |
| • PVOD | 1 (2.9) |
| • CHD | 1 (2.9) |
| Cardiopulmonary Haemodynamics (mean ±SD) | |
| • CI | 2.8 (0.7) |
| • mPAP | 46.2 (13.5) |
| • PVR | 615 (358) |
| • mRAP | 8.6 (4.6) |

**Notes:** Data are presented as numbers (%) for categorical variables, mean (standard deviation) for normally distributed continuous variables, and median (interquartile range) where appropriate. **Abbreviations:** BMI = Body Mass Index; ISWT = Incremental Shuttle Walk Test; WHO = World Health Organisation; NT-proBNP = N-terminal pro-B-type natriuretic peptide; IPAH = Idiopathic Pulmonary Arterial Hypertension; HPAH = Heritable Pulmonary Arterial Hypertension; CTD = Connective Tissue Disease; PVOD = Pulmonary Veno-Occlusive Disease; CHD = Congenital Heart Disease; CI = Cardiac Index; mPAP = Mean Pulmonary Artery Pressure; PVR = Pulmonary Vascular Resistance; mRAP = Mean Right Atrial Pressure.

**Figure 1:**
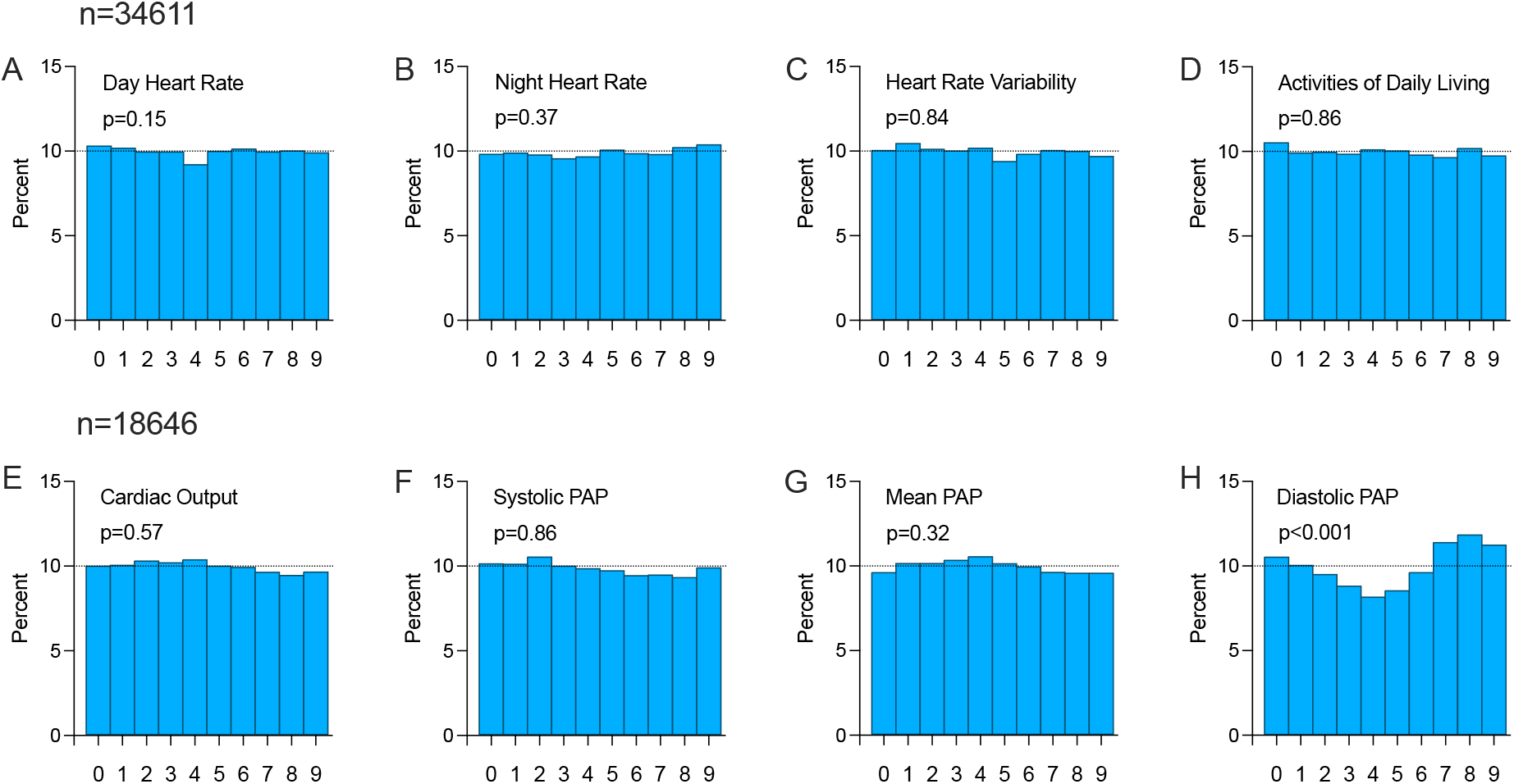
Distribution of terminal digits for (A) day heart rate, (B) night heart rate, (C) heart rate variability, (D) activities of daily living, (E) cardiac output, (F) systolic pulmonary artery pressure, (G) mean pulmonary artery pressure, and (H) diastolic pulmonary artery pressure. The percentage of measurements with each terminal digit is represented on the y-axis, and the terminal digit on the x-axis. P-values are from chi-square tests. n = 34,611 insertable cardiac monitors and n = 18,646 pulmonary artery pressure monitors, derived measurements (follow-up) for PA pressures.

## Discussion

Terminal digit preference is most frequently a function of observer bias. It has been described in studies of patients with systemic hypertension, where it is associated with lower mean pressure and higher numbers of cardiovascular events.^4^ Accurate measures of pulmonary artery pressure, cardiac output, and 6-minute walk distance are critical in the diagnosis and management of patients with pulmonary arterial hypertension, as well as in the evaluation of therapeutic response in clinical studies.^5,6^ However, in phase III randomised clinical trials of PAH therapies, terminal digits 0 and 5 are overrepresented in measures of pulmonary artery pressure and 6-minute walk distance.^3^ In such clinical studies, patients undergo right heart catheterisation before and after administration of a drug, meaning that rounding of values based on terminal digit preference introduces measurement error on two occasions. This non-differential error falsely increases variance, reduces power, and biases studies towards a null outcome. In clinical practice, right heart catheterisation is used for clinical diagnosis and evaluation of treatment response.^7^ In this context, terminal digit preference may alter which patients meet diagnostic criteria and influence ongoing clinical management decisions. As such, methods that provide accurate, unbiased measures of pulmonary artery pressure and cardiac output offer potential advantages. In clinical studies, reduced error may increase study power and reduce the number of patients exposed to drugs and the study duration. In clinical practice, accurate haemodynamic evaluation may ensure clinical decisions are made based on accurate measures. In both circumstances, the use of computer-generated measures and/or core-lab analysis may be preferable to physician-interpreted measures, which are subject to terminal digit preference.^8^

Technological advances have provided the capacity to remotely measure cardiopulmonary haemodynamics and physical activity from the patient’s home. Digital measures of pulmonary artery pressure and physical activity are measured as integers, and cardiac output as decimals.^9^ In contrast to operator-reported values in phase III studies, the present study demonstrates that terminal digit preference was not present in systolic and mean pulmonary artery pressure, cardiac output, heart rate, heart rate variability and physical activity captured using remote, digital technology. A terminal digit preference was observed for 0, 7, 8 and 9 in remote, digitally reported diastolic pulmonary artery pressure. Assuming a uniform distribution of terminal digits should be represented at 10%. The normal range for diastolic pulmonary artery pressure is 8-12 mmHg, and values of less than 5 mmHg are uncommon.^10^ As such, there is a cut-off at the lower physiological range of diastolic pulmonary artery pressure, which biases the distribution of values, meaning the assumption of uniform distribution of terminal digits is not valid for this value.

## Conclusions

We found that digital data collection prevents the described observer-mediated preference for a terminal digit of 0 or 5 in measurements of systolic and mean pulmonary artery pressure in patients with pulmonary arterial hypertension. The minimisation or removal of terminal digit preference bias has important implications for research and clinical care in pulmonary hypertension, in which measures of pulmonary artery pressure, cardiac output and exercise capacity are used for the evaluation of therapeutic efficacy.

## Data Availability

All data produced in the present study are available upon reasonable request to the author

